# Factors associated with psychological distress in health-care workers during an infectious disease outbreak: A rapid living systematic review

**DOI:** 10.1101/2020.07.23.20160879

**Authors:** Fuschia M. Sirois, Janine Owens

**Affiliations:** Department of Psychology, University of Sheffield, United Kingdom; School of Clinical Dentistry, University of Sheffield, United Kingdom

## Abstract

**Background:** Health-care workers (HCW) are at risk for psychological distress during an infectious disease outbreak due to the demands of dealing with a public health emergency.

**Aims:** To examine the factors associated with psychological distress among HCW during an outbreak.

**Method:** We systematically reviewed literature on the factors associated with psychological distress (demographic characteristics, occupational, social, psychological, and infection-related factors) in HCW during an outbreak (COVID-19, SARS, MERS, H1N1, H7N9, Ebola). Four electronic databases were searched (2000 to 10 July 2020) for relevant peer-reviewed research according to a pre-registered protocol. A narrative synthesis was conducted to identify fixed, modifiable, and infection-related factors.

**Results:** From the 3335 records identified, 52 with data from 54,800 HCW were included. All but two studies were cross-sectional. Consistent evidence indicated that being female, a nurse, experiencing stigma, maladaptive coping, having contact or risk for contact with infected patients, and being quarantined, were risk factors for psychological distress among HCW. Personal and organisational social support, perceiving control, positive work attitudes, sufficient information about the outbreak and proper protection, training and resources, were associated with less psychological distress.

**Conclusions:** HCW who may be most at risk for psychological distress during an outbreak require early intervention and ongoing monitoring as there is some evidence that HCW distress can persist for years after an outbreak. Further research is needed to track the associations of risk factors with distress over time and the extent to which certain factors are inter-related and linked to sustained or transient distress.

## Introduction

Several outbreaks of viral diseases have posed significant public health threats since 2000. These include SARS, H1N1, H7N9, MERS, EBOLA, and more recently, COVID-19 (See Supplementary Table 1). Such outbreaks place a serious strain on the health-care systems that try to contain and manage them, including health-care workers (HCW) who are at increased risk for nosocomial infections (1). In addition to the threat to their own physical health, HCW can experience psychological distress as a collateral cost of the risk of infection and the demands of dealing with a public health emergency (2).

Psychological distress refers to a state of emotional suffering, resulting from being exposed to a stressful event that poses a threat to one’s physical or mental health (3). Inability to cope effectively with the stressor results in psychological distress that can manifest as a range of adverse mental health and psychiatric outcomes including depression, anxiety, acute stress, post-traumatic stress, burnout, and psychiatric morbidity. Although psychological distress is often viewed as a transient state that negatively impacts day-to-day and social functioning, it can persist and have longer-term negative effects on mental health (4).

Under normal circumstances, work-related psychological distress in HCW is associated with several short and long-term adverse outcomes. Psychological distress is linked to adverse occupational outcomes including include decreased quality of patient care (5), irritability with colleagues (6), cognitive impairments that negatively impact patient care (7), and intentions to leave one’s job (8). HCW who experience psychological distress are also at risk of experiencing adverse personal outcomes including substance misuse (6), and suicide (9). In the context of an infectious disease outbreak, such consequences are likely amplified to the extent that psychological distress is heightened. HCW who reported elevated levels of psychological distress during the COVID-19 outbreak also experienced sleep disturbances (10), poorer physical health (11), and a greater number of physical symptoms, including headaches (12). Similarly, HCW during the SARS outbreak disclosed a greater number of somatic symptoms and sleep problems (13), substance misuse and more days off work (14).

HCW serve a vital role in treating and managing infected individuals during an outbreak. Given the ongoing coronavirus outbreak, there is an urgent need to understand the factors that create or heighten risk for distress for HCW and impact their immediate and long-term mental health, as well as those that are protective and may reduce psychological distress. Such knowledge is important for identifying HCW most at risk, and informing strategies and treatments needed to support HCW resilience during and after an outbreak.

This rapid review synthesised the evidence on the factors associated with psychological distress among health-care workers (HCW) during an infectious disease outbreak. It also identified and classified the factors that contributed to risk or provided resilience for psychological distress. The key questions were:

1. What are the risk factors for psychological distress among HCW during an infectious outbreak?
2. What are the factors associated with reduced risk for psychological distress among HCW during an infectious outbreak?

## Methods

Evidence was summarised using a rapid, living review approach because of the urgent need to support the mental health of HCW during and after the ongoing novel coronavirus pandemic. Rapid Reviews provide an expedient and useful means of synthesising the available evidence during times of health crises to inform evidence-based decision making for health policy and practice (15, 16). To accomplish this, rapid reviews take a streamlined approach to systematically reviewing evidence. Modified methods in the current review included: 1) search limited to English language studies; 2) grey literature limited to one search source; 3) no formal critical appraisal of the research. Living reviews feature continual updating of the review as new evidence is released (17).

### Data Sources and Searches

The search strategy for this pre-registered rapid review involved searching Medline, PsychInfo, Web of Science, and the first 10 pages of Google Scholar, as well as hand searching references. Search terms included a combination of terms related to health-care workers (e.g., “physicians”, “nurses”), and distress (e.g., “stress”, “anxiety”). The full search term list is available on PROSPERO (CRD42020178185). We conducted searches in a rolling manner, starting on April 6, 2020, then with updates on June 7, July 2, and July 10, 2020 to capture and integrate the most up-to-date evidence given the ongoing COVID-19 outbreak and the associated rapid release of research. Searches for new studies is ongoing and will use the same search methods described above Should new evidence be found that substantially changes the conclusions, a major update will be performed. Otherwise, new evidence will be updated monthly for a 6-month period.

### Study selection and data extraction

A predefined search strategy was used (see full details on PROSPERO, https://www.crd.york.ac.uk/PROSPERO/; registration ID: CRD42020178185). Studies were included in this Review if they were empirical research; published or accepted for publication in peer-reviewed journals; written in English; included participants who were HCW in a hospital environment during a major infectious outbreak (COVID19, SARS, MERS, H1N1, H7N9, Ebola); had a sample size of greater than 80, and included data on factors associated with psychological distress during an outbreak. One investigator screened citations for potential full-text review, and a second investigator conducted the full-text review of each study for inclusion. Exclusions were verified by another reviewer, and disagreements resolved through discussion. Data was extracted by one reviewer, entered into a table, and verified by a second reviewer.

As this was a rapid review, a formal assessment of study quality and risk for bias was not conducted (15, 16). In lieu of a formal assessment, we only included studies that reported findings for a sample size of greater than 80, which allows enough power to detect a medium effect size with an alpha of 0.05 (18, 19).

### Data Synthesis and Analysis

We conceptually organised the factors in this Review identified as contributing to or mitigating psychological distress into three broad categories: 1) fixed or unchangeable factors (sociodemographic and occupational factors), 2) potentially modifiable factors (social and psychological factors), and 3) factors related to infection exposure. Fixed factors identify which HCW might be most vulnerable or resilient to distress, whereas modifiable factors identify potential targets for interventions to reduce risk and increase resilience. Infection-related factors are those that can directly inform hospital procedures and operating policy regarding ways to address and mitigate risk.

## Results

The search yielded 3334 records, with 52 papers reporting 53 studies (Total *N* = 54,800 HCW) that met inclusion criteria for this Review. Figure 1 presents the complete screening process. Characteristics of the studies are in Table 1. The average sample size was 1,033 (range 82 – 14,825). The studies included HCW working across 15 countries during SARS (21), MERS (7), H1N1 (2), COVID-19 (21), Ebola (1), and H7N9 (1), outbreaks. The rates of psychological distress in HCW varied depending on how distress was measured (Table 1).

**Table 2:**
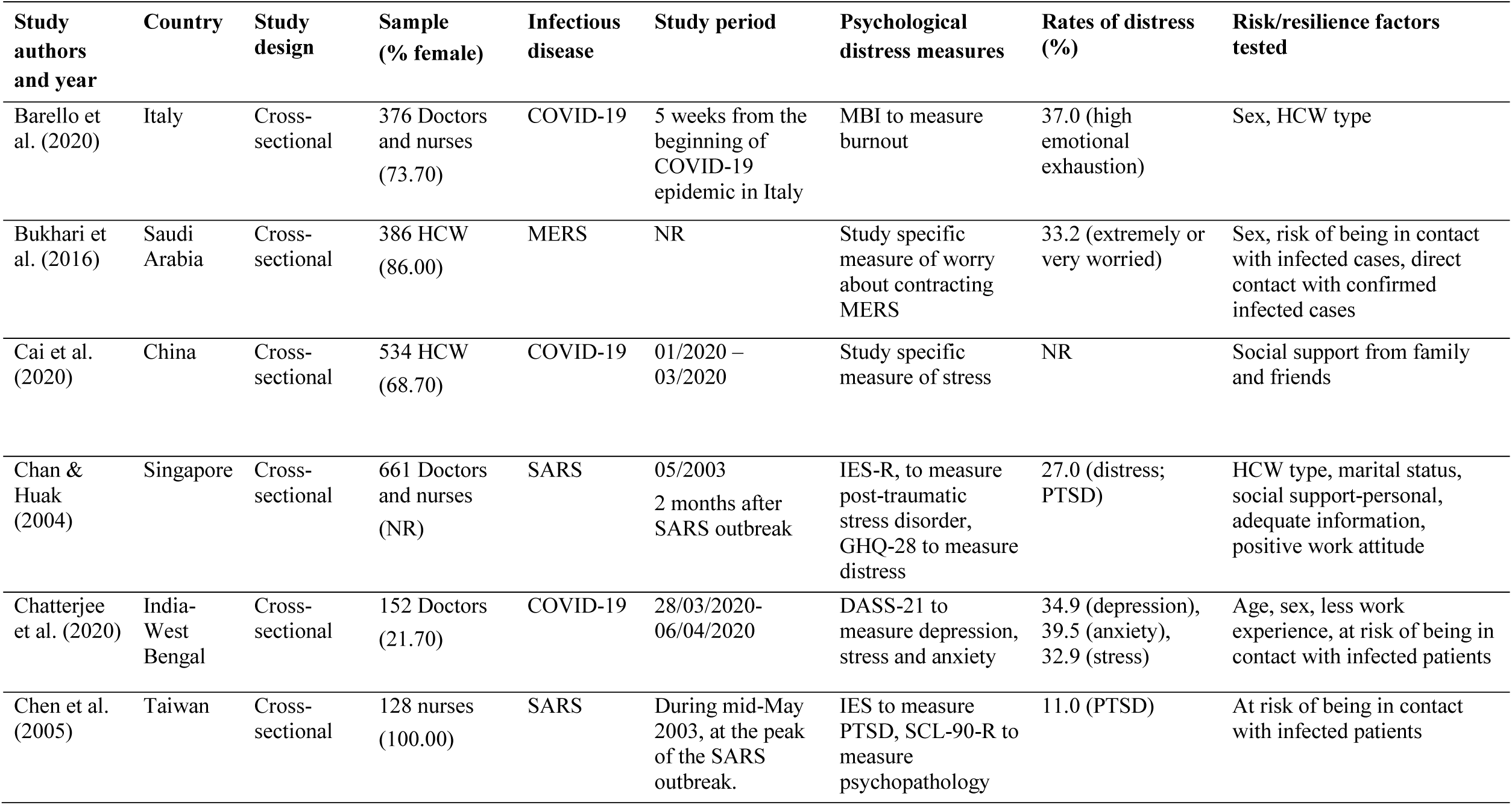
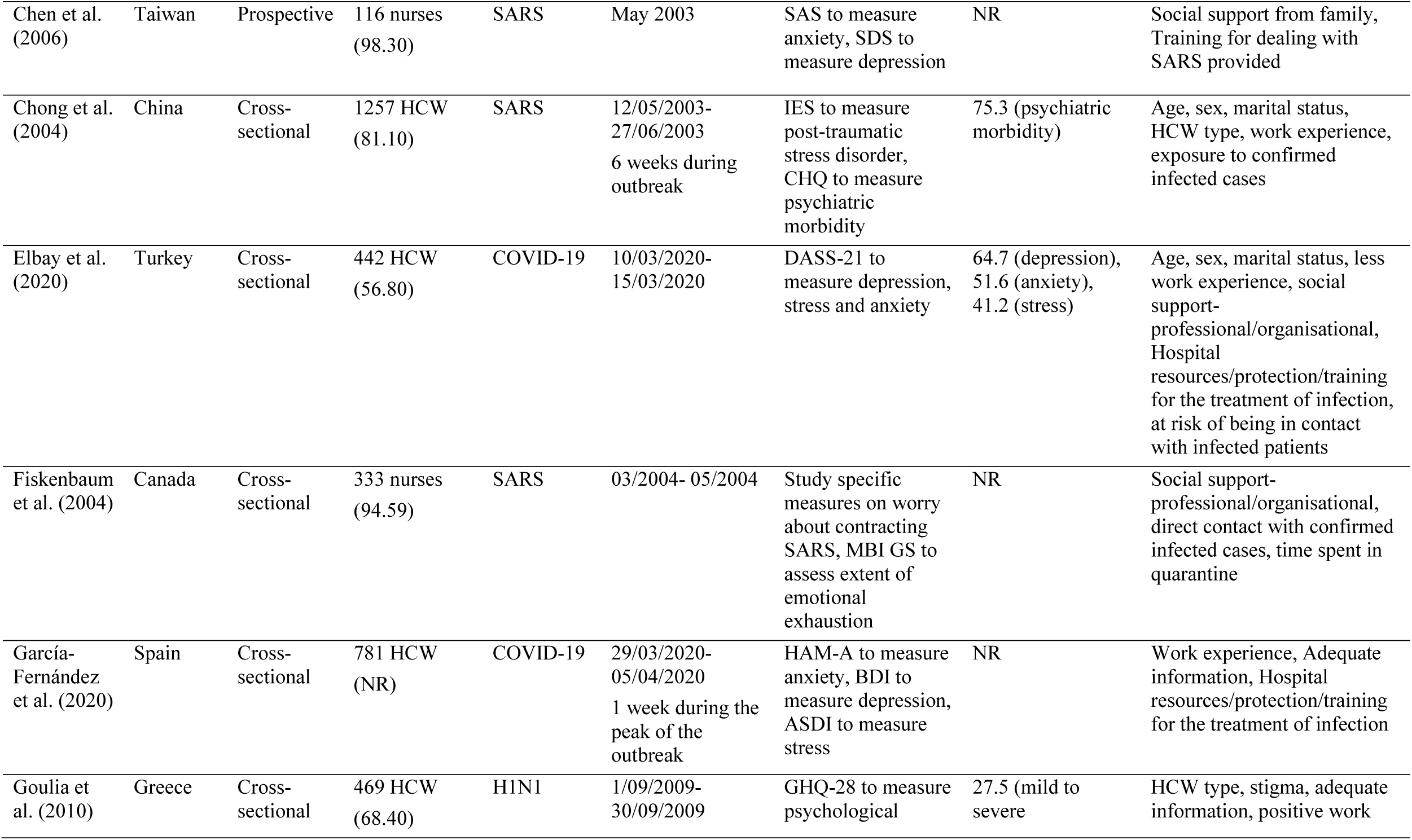
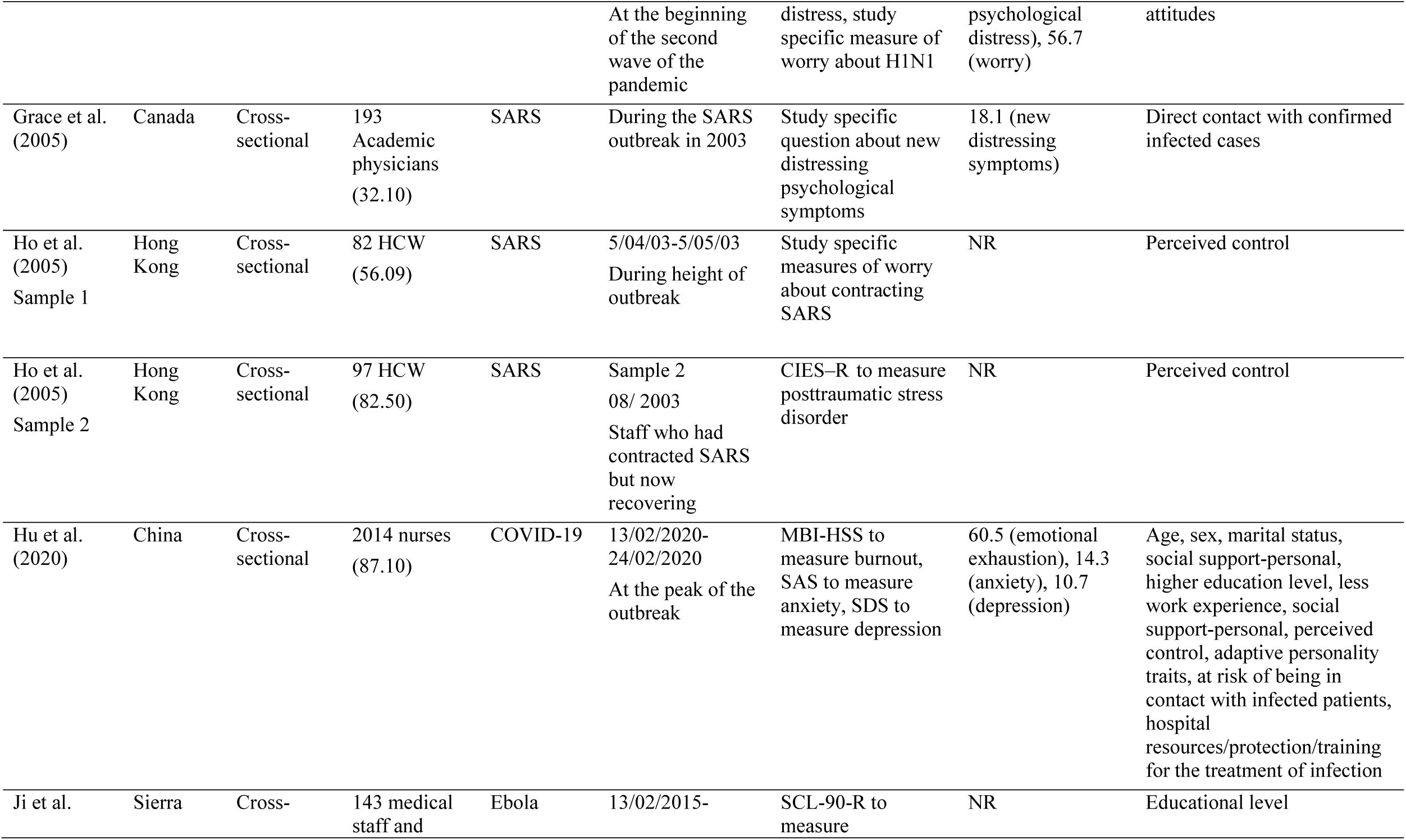
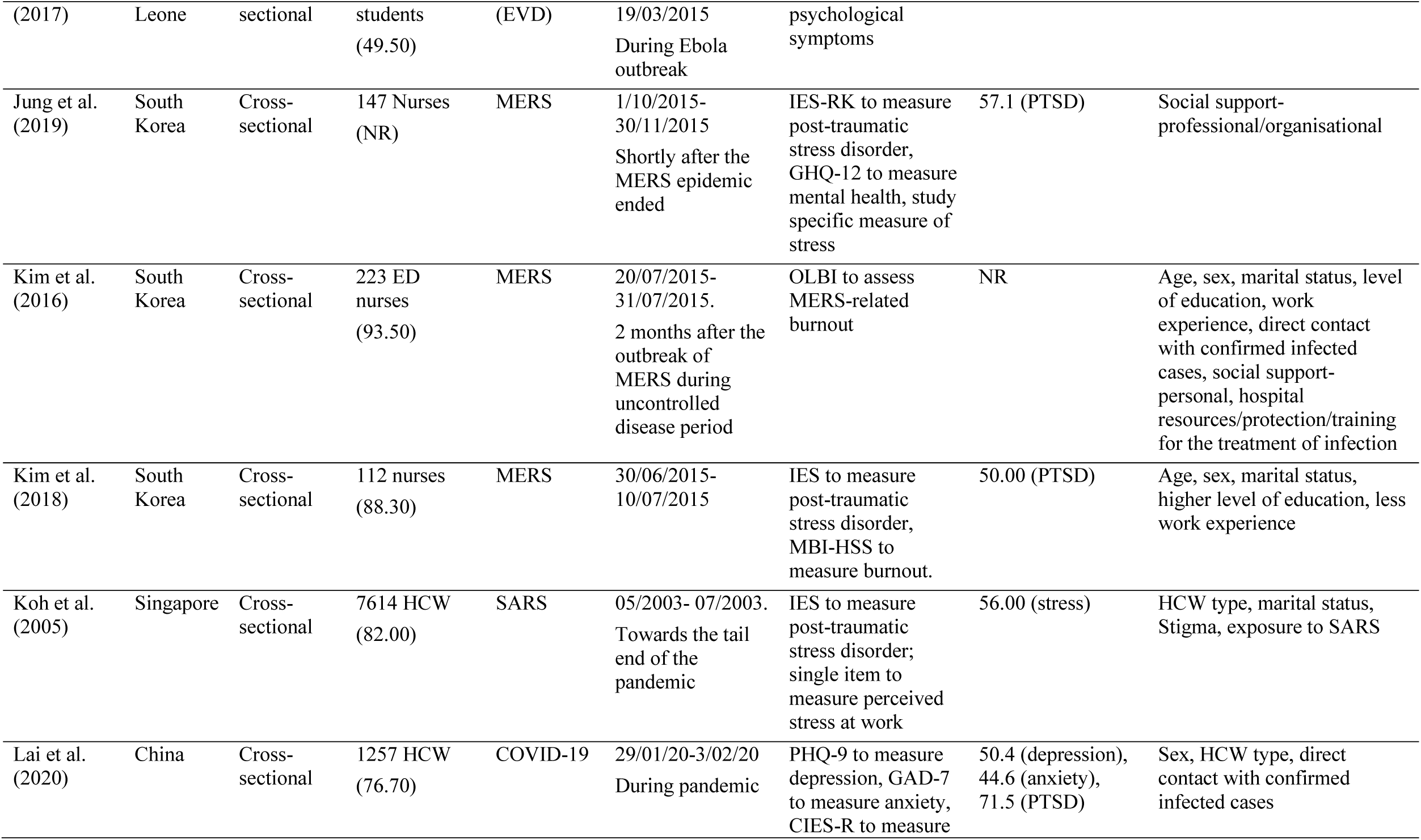
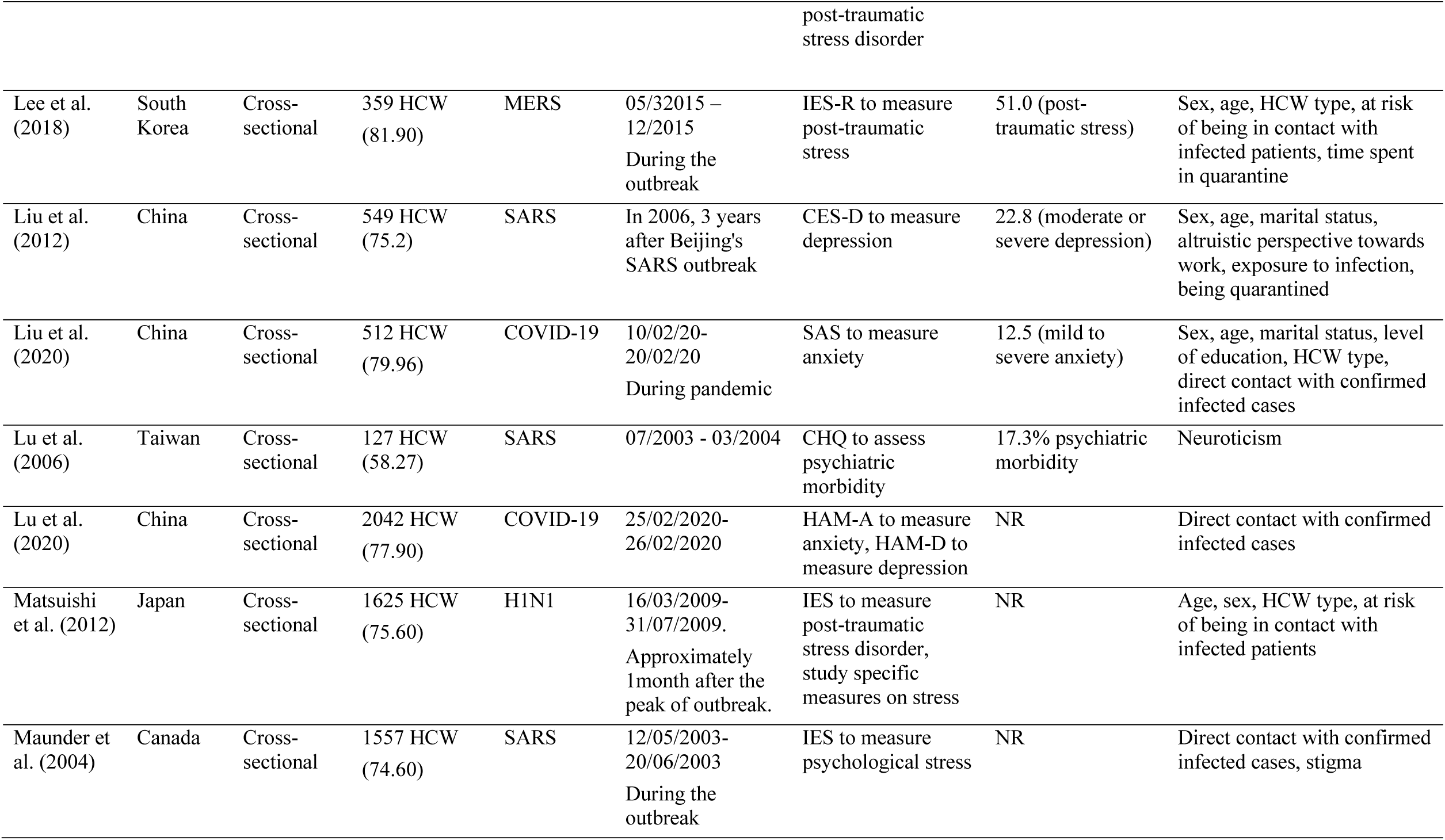
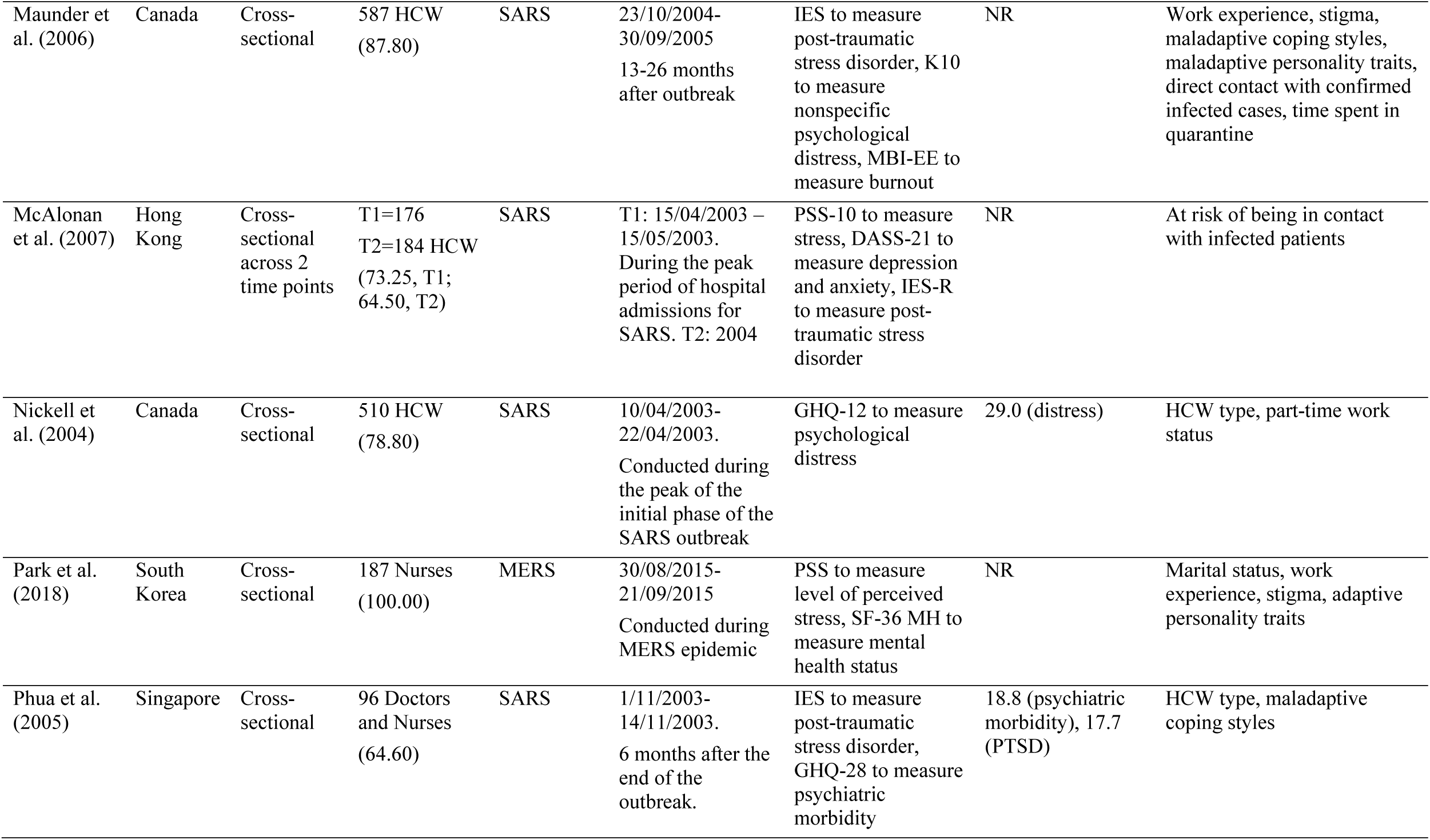
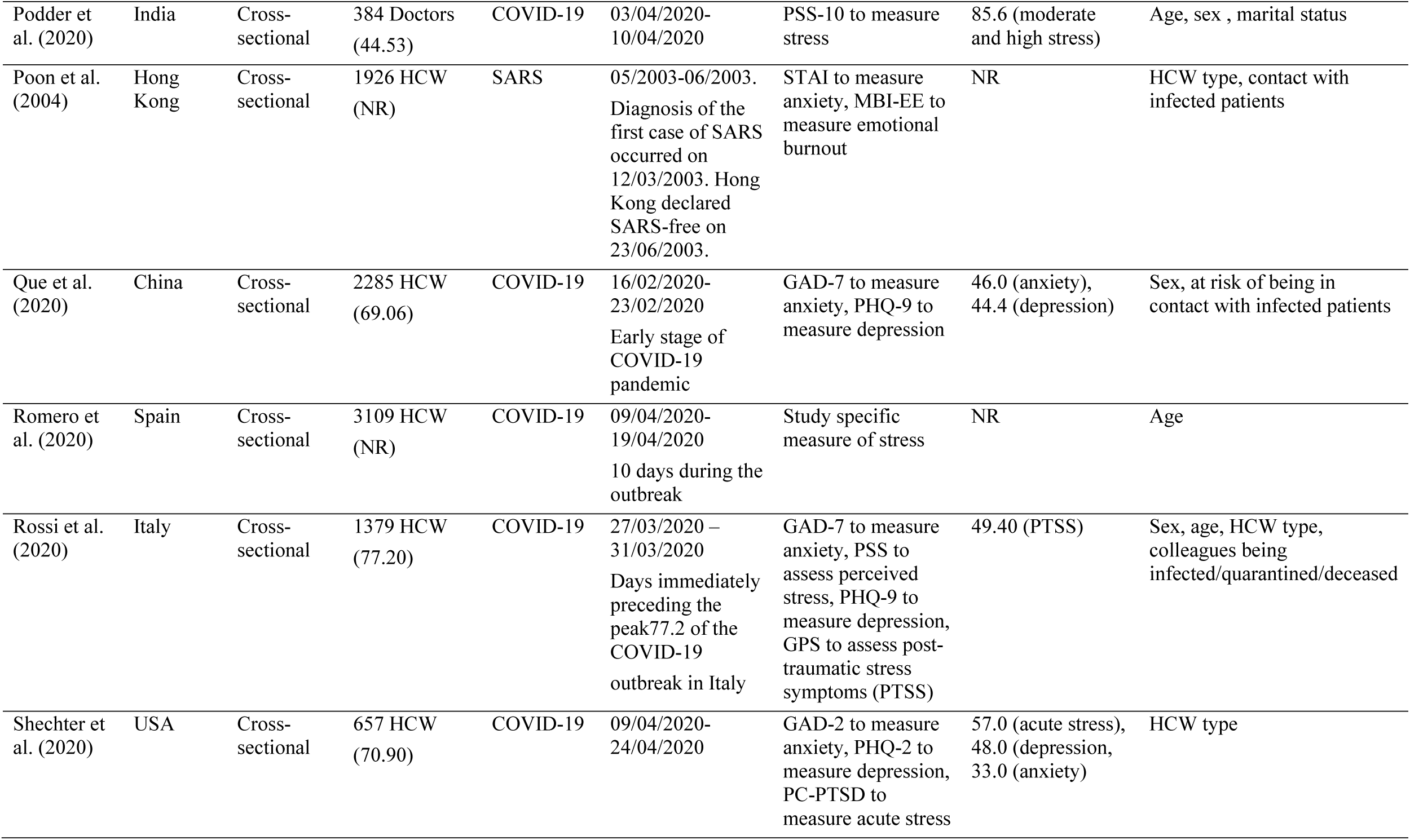
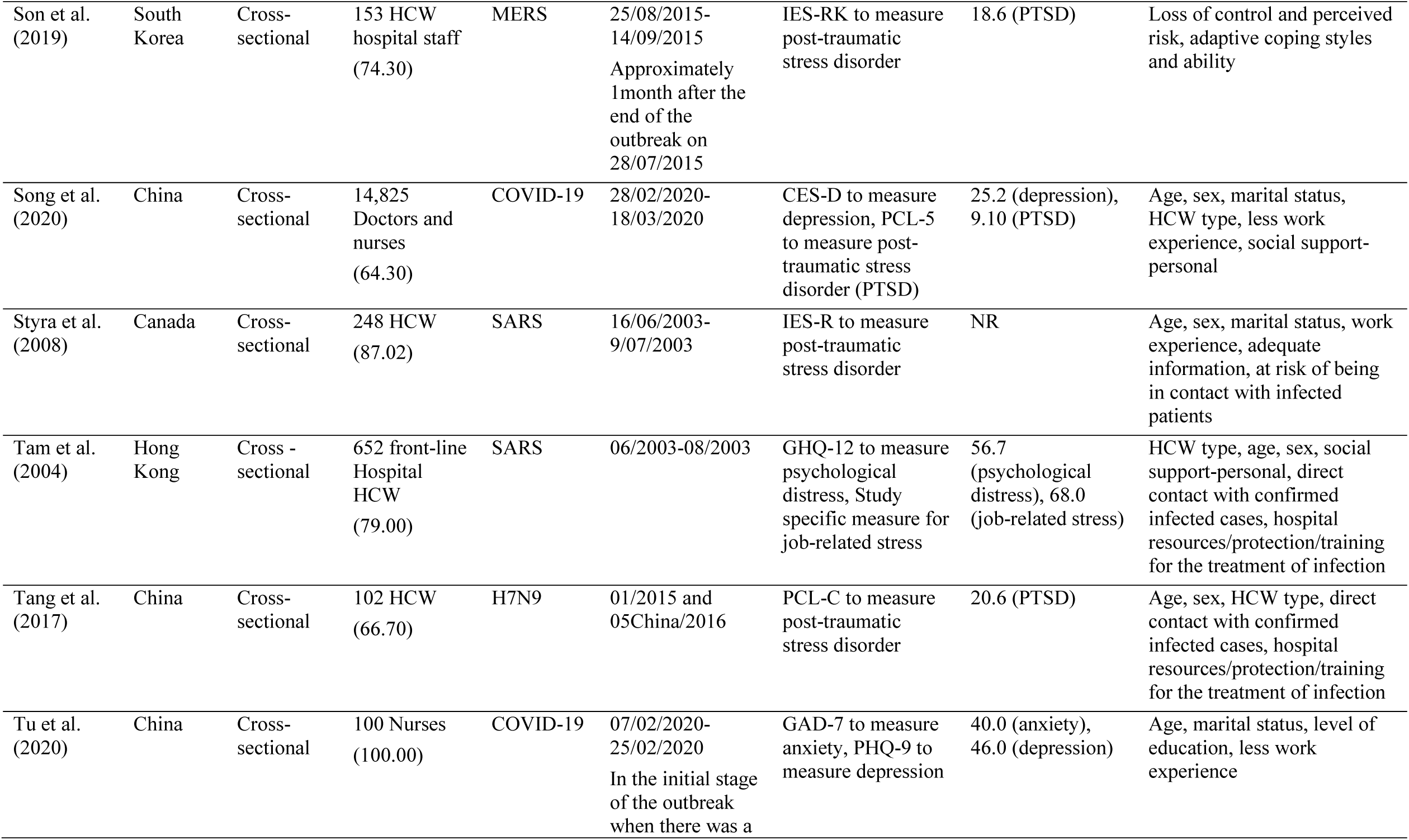
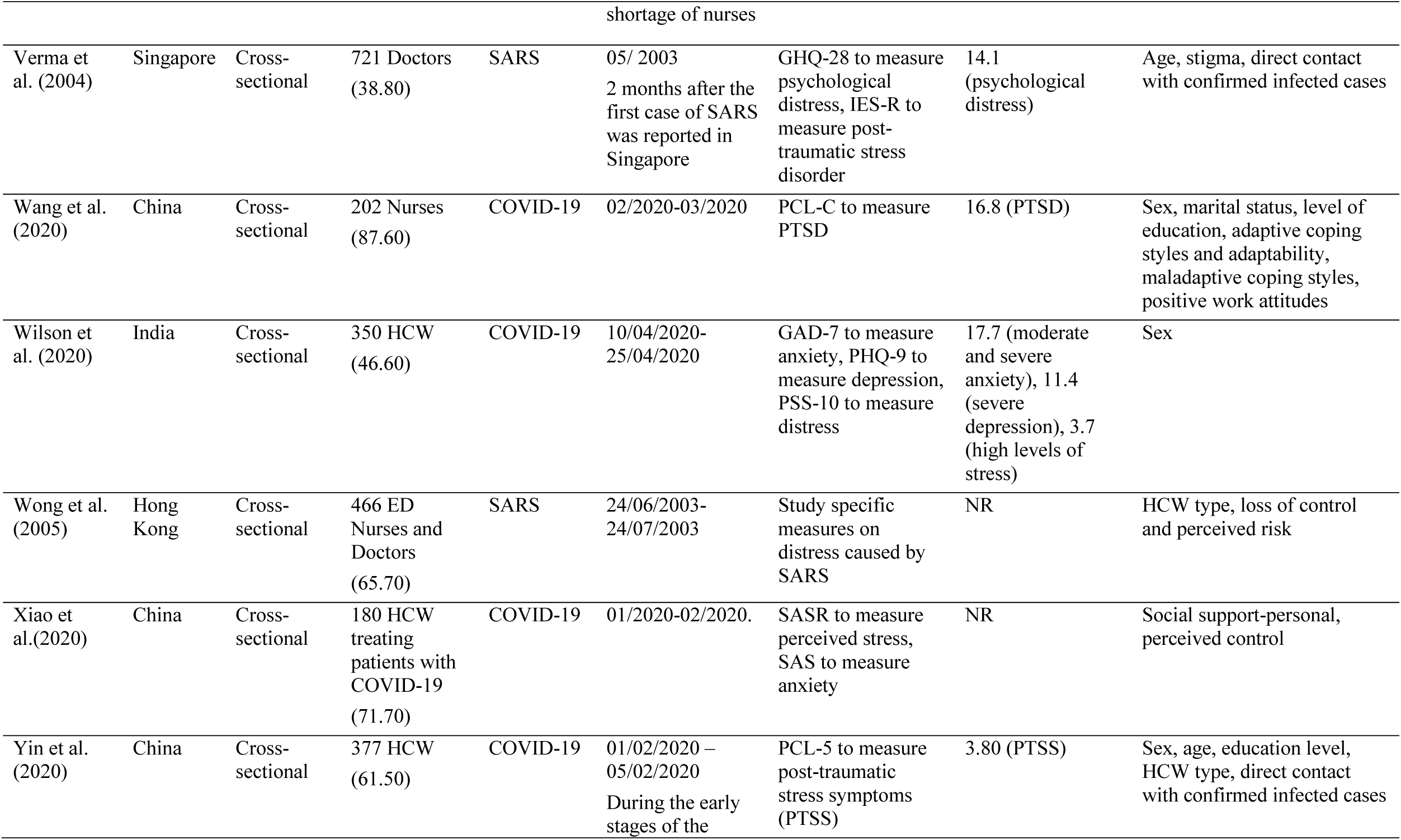
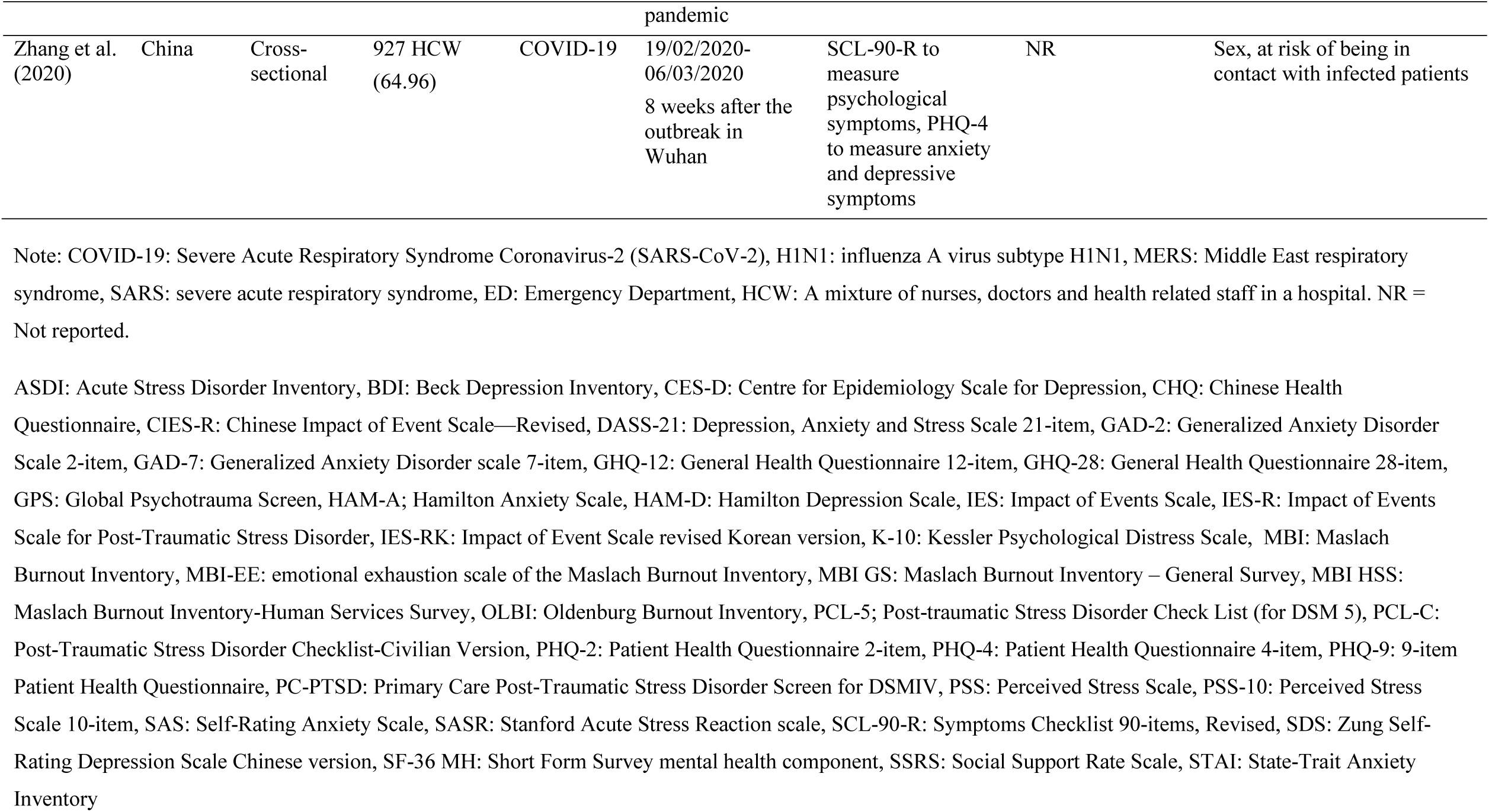
Characteristics of the 53 studies (N = 54,800) included in the rapid review

**Figure 1:**
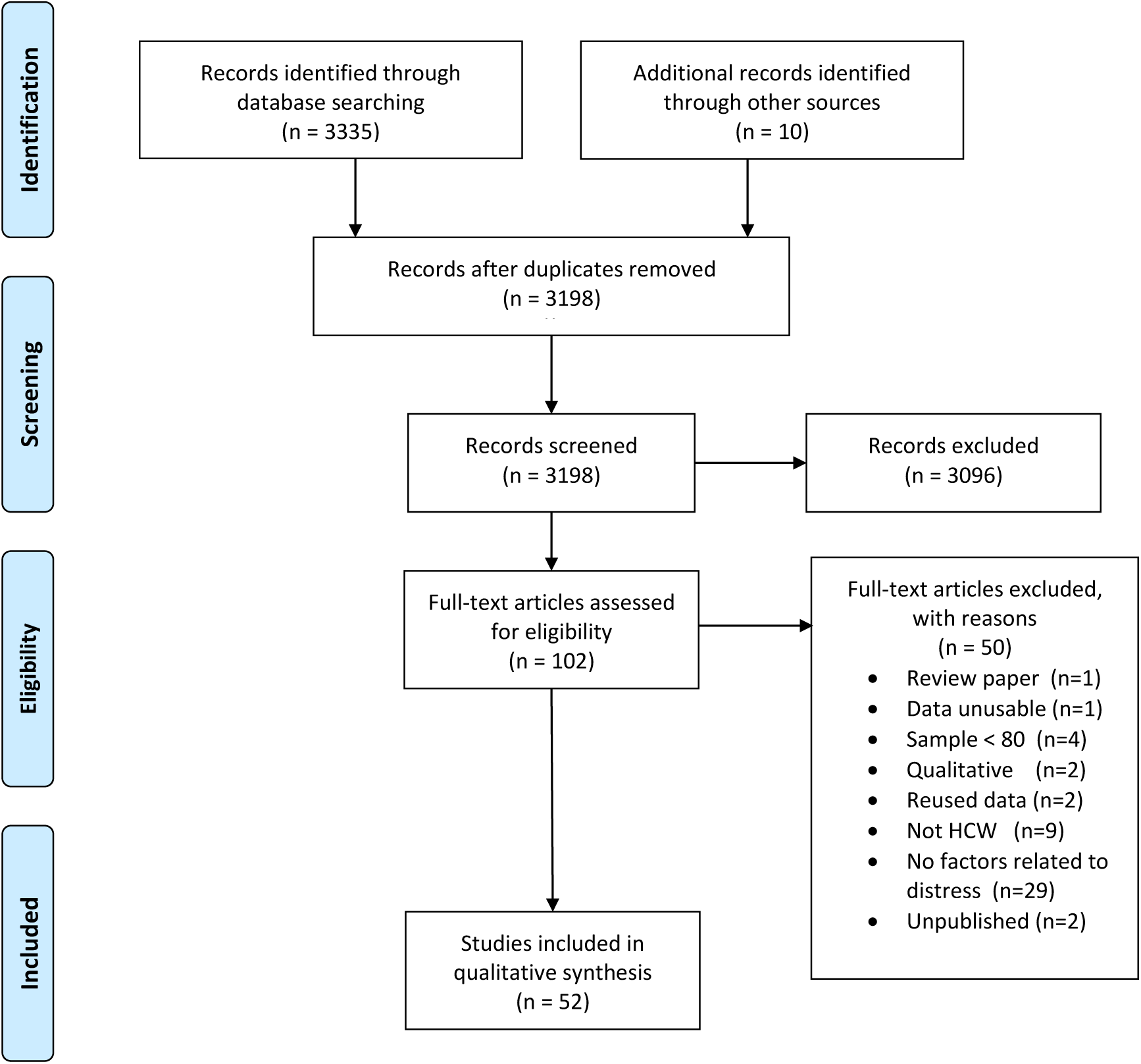
PRISMA flow diagram for literature screening

### Sociodemographic factors

Twenty studies examined age as a predictor of psychological distress among HCW during an epidemic (see Table 2). Of these, ten found that age was a significant risk factor for distress. In two studies of HCW during the SARS outbreak, staff who were younger than 33 experienced greater stress, but not greater psychiatric morbidity, compared to older staff (20), and staff under 35 were more likely to report severe depressive symptoms three years after the outbreak (21). In another study, medical staff who were between 20 and 30 years old and exposed to patients with H7N9 had elevated post-traumatic stress disorder scores compared to older staff (22). Similarly, general practitioners in working during the SARS outbreak who met psychiatric caseness for PTSD were more likely to be younger (23). In a study during the H1N1 outbreak, hospital staff who were in their 20’s had greater anxiety about becoming infected than did older staff (24). During COVID-19, HCW who were younger were more likely to experience higher levels of post-traumatic stress symptoms, depression, anxiety, and acute stress (25-29). In contrast, ten studies found that age was not a significant predictor of distress in HCW during the SARS, MERS or during the COVID-19 outbreaks (Table 2).

**Table 2:**
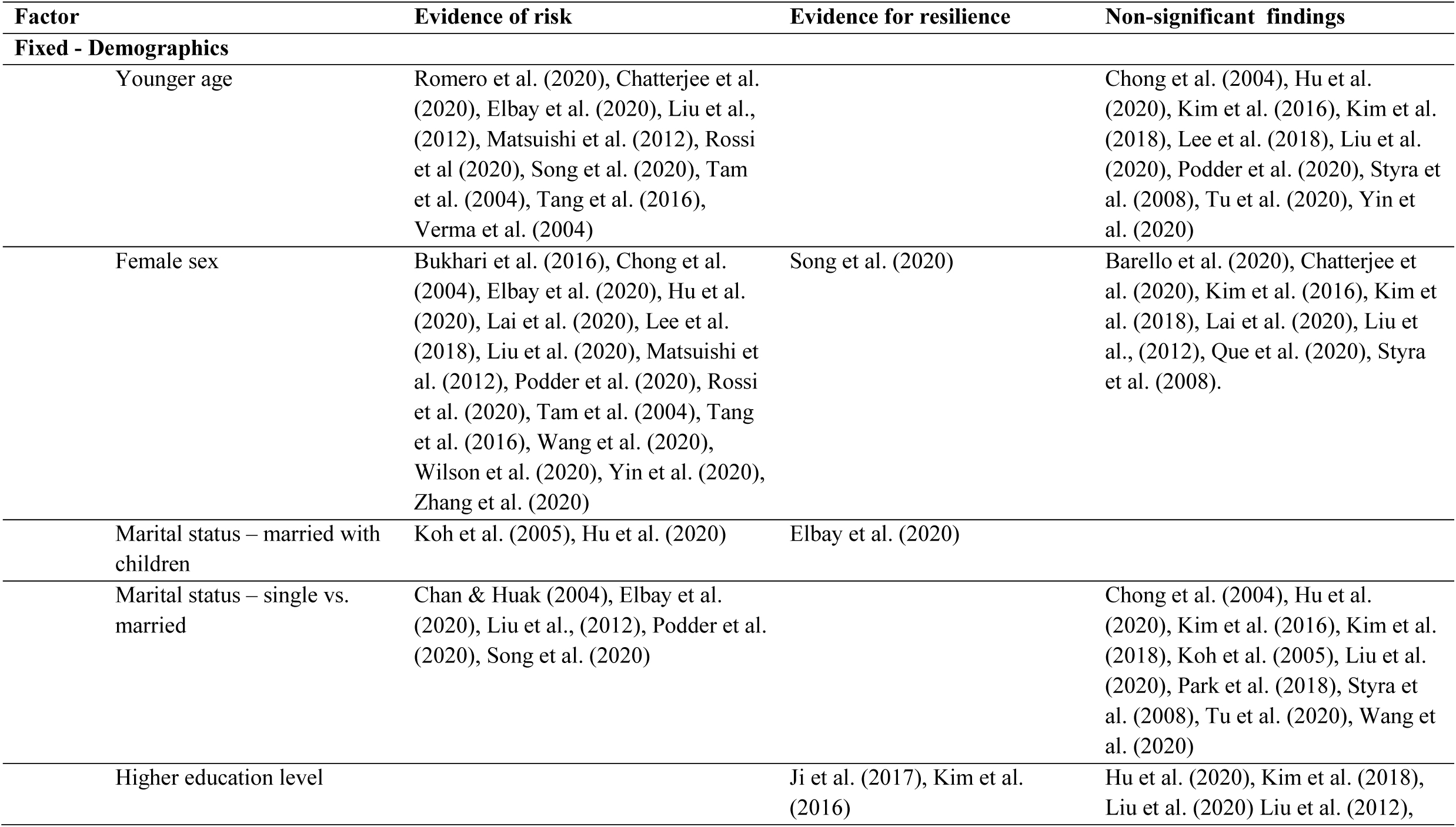
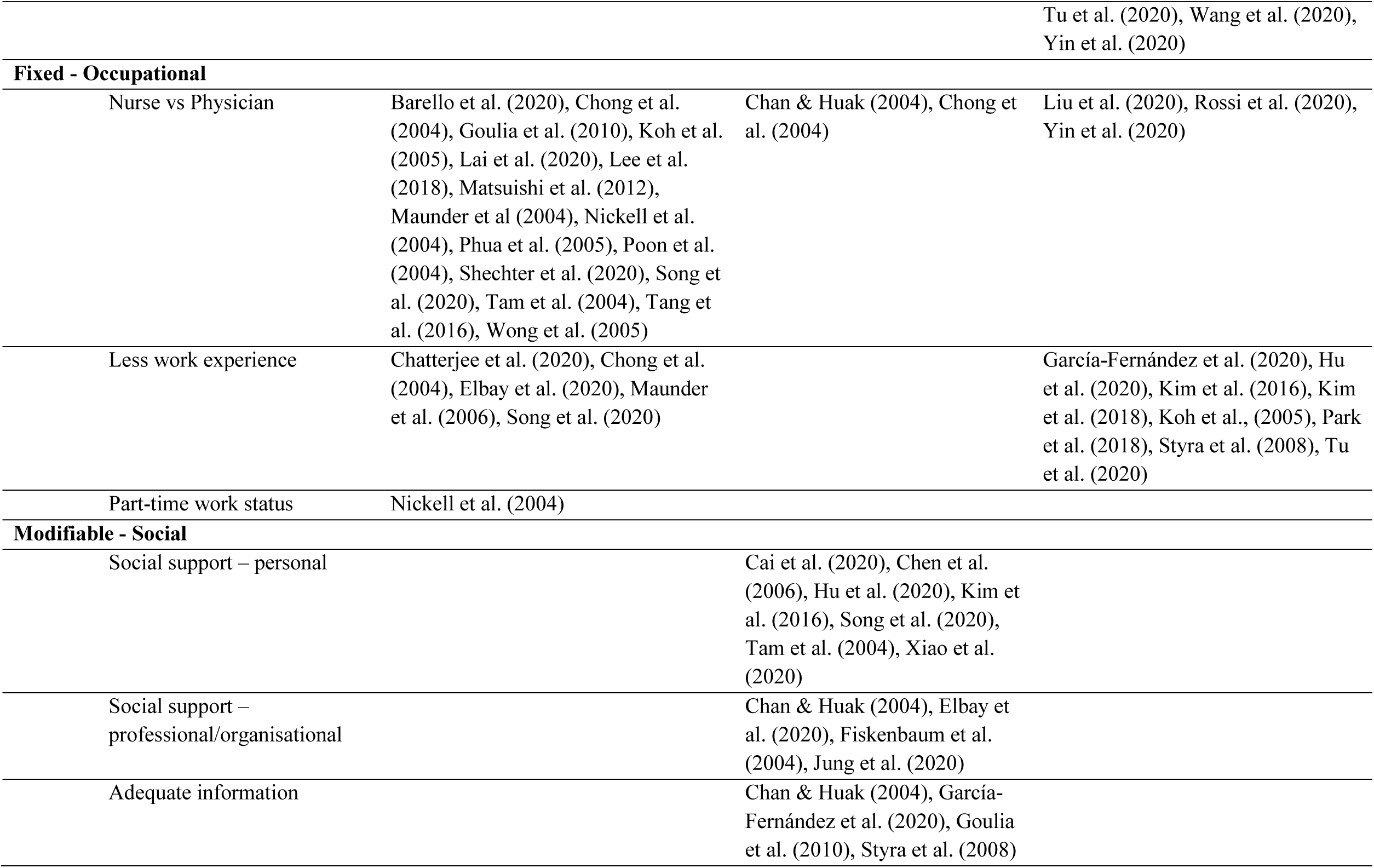
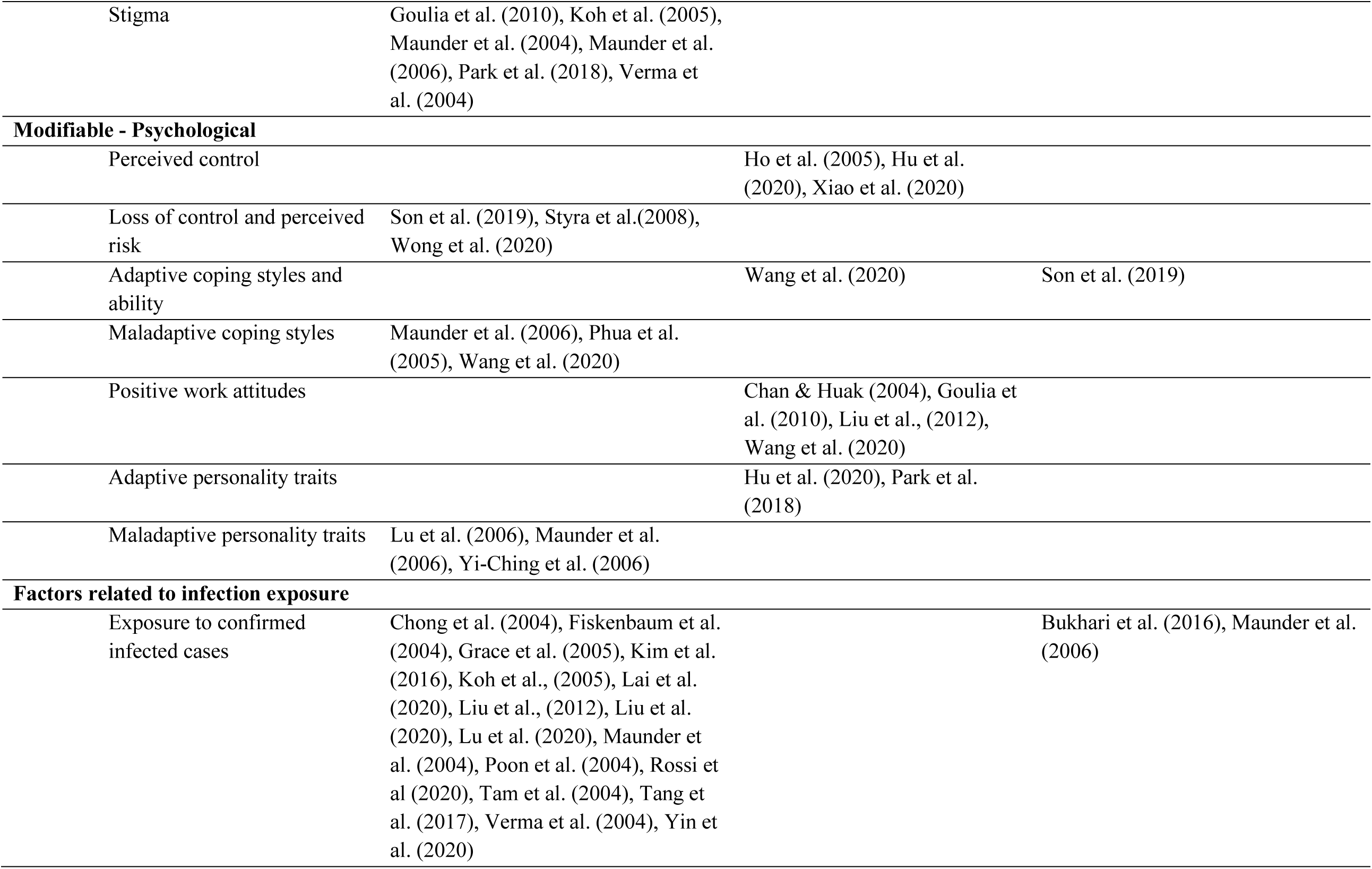
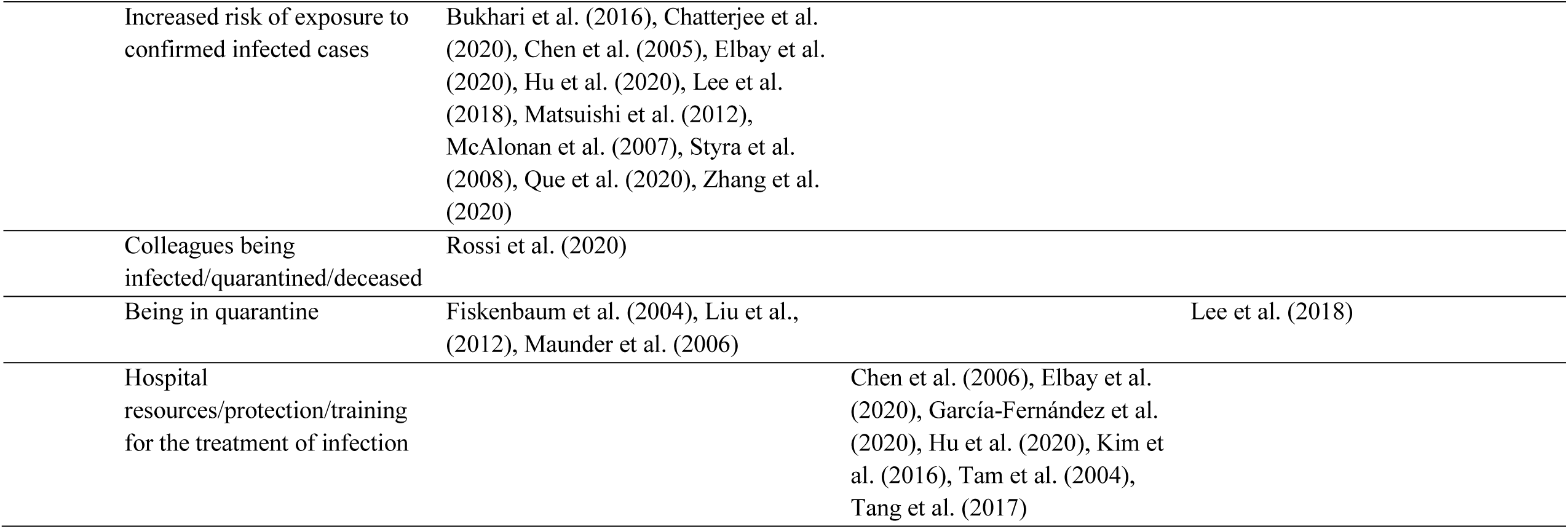
Overview of the evidence for the factors associated with risk and resilience for psychological distress in HCW

Twenty-five studies tested sex as a possible risk factor for distress among HCW during an outbreak (Table 2), with all but nine finding that being female was associated with higher risk for psychological distress. Notably, the fifteen studies that found that female sex was a significant risk factor spanned six different infectious diseases (MERS, SARS, COVID-19, H1N1, H7N9, and SARS), suggesting that being a female HCW increases vulnerability for distress more generally when working during an infectious outbreak.

Of the fifteen studies that examined marital status as a risk or resilience factor for psychological distress, only five found evidence to suggest this as a risk factor (Table 2). Two studies of HCW during the SARS outbreak found that HCW who were single were 1.4 times more likely to experience psychological distress than married HCW (30), and more likely to have sever depressive symptoms three years later (21). Similarly, HCW during the COVID-19 outbreak who were single experienced higher levels of distress than those who were married (27, 31, 32). Conversely, two studies found that married HCW with children reported greater stress than single HCW or those who were married without children (33, 34). Ten other studies conducted during the SARS, MERS, and COVID-19 outbreaks found no associations between HCW marital status and distress.

Nine studies examined education levels in association with distress. Only two studies, conducted during the Ebola outbreak (35), and the MERS outbreak (36) found that HCW with higher educational levels reported significantly lower psychological distress. Education level was not predictive of psychological distress among HCW working during the MERS or the COVID-19 outbreaks (Table 2).

### Occupational factors

Twenty-one studies examined and found evidence that the HCW occupational role created risk for psychological distress while working during the SARS, H1N1, MERS, and COVID-19 outbreaks (Table 2). In all but five studies, being a nurse was associated with a range of mental health issues, including higher stress, burnout, anxiety, depression, PTSD symptoms, psychiatric morbidity, and psychological distress compared to being a physician or other HCW (see Tables 1-2. The extent to which nurses experienced greater psychological distress whilst working during an outbreak was estimated in four studies; nurses were 1.2 (33), 1.4 (37), 2.2 (38), and 2.8 (39) times more likely to be at risk for poor mental health. In contrast, two studies found that physicians (13) and technicians (30) were more likely to experience distress while working during the SARS outbreak. Three studies conducted during the COVID-19 outbreak did not find that occupational role was a risk factor for distress (Table 2).

Other occupational factors examined included years of work experience, and full-time versus part-time status. Only five of the thirteen studies found evidence to suggest that less work experience may create risk (Table 2). HCW who had worked for less than two years experienced significantly greater stress than those with more work experience in a large sample of HCW during the SARS pandemic (13). In HCW during the SARS outbreak, those with less than 10 years of experience reported higher levels of psychological distress, but not burnout or posttraumatic stress, 13-26 months after the outbreak (14). HCW who had less clinical experience were also more likely to experience stress during the COVID-19 outbreak (Table 2). Years of clinical experience was not associated with PTSD symptoms, acute stress or anxiety, depression, mental health status, or burnout in five other studies (Table 2). Lastly, in one study, part-time worker status was a significant predictor of greater emotional distress in HCW during the SARS outbreak (39).

### Social factors

A number of social and interpersonal factors mitigated or contributed to psychological distress. Receiving direct social support from friends, family, colleagues and supervisors was a key protective factor in the eleven studies that examined its association with psychological distress (Table 2). In HCW during the COVID-19 outbreak, higher levels of social support were associated with significantly lower levels of stress, depression, anxiety, depression and PTSD. These findings were consistent with that of a study of frontline medical staff during the COVID-19 outbreak who reported that a positive attitude from co-workers was important for reducing their distress (40). Analogously, emergency nurses working during MERS outbreak who reported poor support from family and friends experienced higher levels of burnout (36). Similarly, studies of HCW during the SARS outbreak found that higher levels of family support was associated with lower depression and anxiety whereas inadequate support from relatives, lack of gratitude from patients and relatives, and perceiving less of a team spirit at work was associated with higher levels of psychological distress (Table 2).

Organisational support was an important factor in buffering psychological distress of HCW during an outbreak. In nurses working during the SARS outbreak in Canada, higher perceived organisational support in the form of receiving positive performance feedback from doctors and co-workers, was associated with lower perceptions of SARS-related threat and reduced feelings of emotional exhaustion (41). Similarly, nurses, physicians, and HCW working during the MERS, COVID-19 and SARS outbreaks who perceived support from their supervisors and colleagues, experienced better mental health in the form of lower PTSD symptoms, lower distress, and being less likely to develop psychiatric symptoms, respectively (Table 2).

Four studies examined receiving useful information from others (a common form of social support). In one study, HCW who received adequate communication and information about the H1N1 outbreak from their organisation were less likely to experience psychiatric symptoms because it helped them cope better, and worry less about the pandemic (38). Similarly, HCW during the SARS outbreak who had confidence in the information they received from their organisation (42), and who received clear communication about directives and how to take precautionary measures (30), experienced reduced psychological distress. HCW working during the COVID-19 outbreak who felt that they did not receive sufficient information, scored significantly higher on anxiety and acute stress than those who were satisfied with the information provided (43).

Negative social perceptions created risk for poor mental health for HCW in six studies. In nurses during the MERS outbreak, perceived social stigma was associated with higher stress and poorer mental health (44). Similarly, general physicians during the SARS outbreak (23), who perceived stigma concerning negative public attitudes and disclosing about one’s work, or self-stigma, experienced higher psychological distress. During the SARS outbreak, HCW who felt people avoided their family because of their job were twice as likely to have elevated levels of post-traumatic stress symptoms (33). Importantly, experiencing stigma and avoidance from others was significantly associated with higher levels of posttraumatic stress symptoms during the SARS outbreak (45), and 13-26 months later (14).

### Psychological factors

The psychological factors examined in the studies included adaptive and maladaptive coping responses, beliefs and attitudes, and personality traits. Six studies examined how perceptions of control were associated with distress among HCW (Table 2). In three studies, higher self-efficacy, was associated with lower anxiety, depression, distress, and lower levels of fear about SARS and post-traumatic stress symptoms during the COVID-19 and SARS outbreaks, respectively. Conversely, feeling a loss of control was associated with greater distress (46) during the SARS outbreak in Hong Kong. Analogously, appraisals of personal risk were linked to higher levels of PTSD symptoms in HCW during the MERS (32) and SARS (42) outbreaks.

Positive attitudes towards one’s work were protective against distress in four studies. Higher work satisfaction was associated with less psychological distress among hospital staff during the H1N1 outbreak (38), and lower PTSD among nurses during the COVID-19 outbreak (47). Similarly, HCW during the SARS outbreak who felt their work had become more important were less likely to develop psychiatric symptoms (30), and those who viewed their work altruistically were less likely to have severe symptoms of depression 3 years later (21).

Five studies examined whether coping styles were associated with HCW distress during an outbreak. Emergency physicians and nurses working during the SARS outbreak who used denial, mental disengagement, or venting of emotions to cope were more likely to score higher on psychiatric morbidity (48). Similar results were found in a study of nurses exposed to COVID-19, with use of negative coping associated with higher PTSD and positive coping linked to lower PTSD (47). In HCW during the SARS outbreak, those who used maladaptive coping strategies, such as escape-avoidance, and self-blame coping, reported higher levels of burnout, psychological distress, and post-traumatic stress when surveyed 13-26 months after the outbreak (14). However, the use of adaptive strategies, such as problem-solving and positive reappraisal, were not associated with any of the distress outcomes. This finding was consistent with a study in which coping ability was not significantly associated with PTSD symptoms during the MERS outbreak (32).

Five studies investigated the role of personality in psychological distress. During the SARS outbreak, neuroticism was linked to poorer mental health (49), and HCW who had an anxious attachment style reported experiencing higher burnout, psychological distress, and posttraumatic stress 13-26 months after the outbreak (14). Those with an avoidant attachment style reported greater distress, but not burnout or posttraumatic stress. Two studies examined the role of dispositional resilience. Among nurses working during the MERS outbreak, higher levels of hardiness were associated with lower stress and better mental health (44), and resilience was associated with lower anxiety, depression, and burnout among frontline nurses during COVID-19 (34).

### Factors related to infection exposure

Eighteen studies examined the impact of direct contact with infected patients on HCW’s psychological distress. Of these, fourteen found that being in direct contact with and/or treating patients infected with COVID-19, SARS, MERS or H7N9 was a risk factor for psychological distress (Table 2). Only two studies did not find that contact with infected patients increased risk for distress (14, 50). Similarly, eleven studies found that risk of contact with infected patients due to working in high-risk areas (e.g., ICU, isolation areas and infection units) was associated with higher levels of anxiety, stress, and post-traumatic stress symptoms than not working in such areas (Table 2). Notably, one study found that HCW in a high-risk unit during SARS reported higher and sustained perceived stress one year after the outbreak compared to those in low-risk units, with those in low-risk units reporting a decrease in stress over time, but those in high-risk units experiencing an increase in stress post-outbreak (51). Spending time in quarantine due to risk of being infected was associated with higher levels of burnout, depression, and psychological distress in HCW during SARS, but was unrelated to post-traumatic stress symptoms in HCW during the MERS outbreak (Table 2). Lastly, one study found that HCW who had colleagues who became infected, had deceased due to infection, or had been quarantined, also experienced higher levels of post-traumatic stress symptoms and acute stress during the COVID-19 outbreak (25).

Provision of adequate training, protection and other resources to manage and reduce risk of infection was associated with less psychological distress in seven studies. Receiving clear infection control guidelines predicted lower psychological morbidity in frontline HCW during SARS (20), and having sufficient hospital resources for the treatment of MERS was associated with lower MERS-related burnout (36). After the implementation of a SARS protection training program, HCW experienced significant decreases in anxiety and depression two weeks and one month after the starting the program (52). Similarly, medical staff receiving inadequate training related to managing H7N9 had higher PTSD symptoms than those who received appropriate training (36). During COVID-19, HCW who felt that the protection they were given was insufficient, felt unsafe, and perceived lower logistic support, reported higher levels of depression, anxiety, and acute stress symptoms (Table 2).

## Discussion

This rapid living systematic review identified three main categories of factors contributing to increased and reduced risk of psychological distress among HCW during an infectious disease outbreak. For the fixed factors (demographic and occupational), the weight of the evidence indicated that HCW who were female or a nurse were at significant risk for psychological distress (Figure 2). Nurses tend to tend to be predominantly female, have higher workloads (45), and have more patient contact than other HCW. There was also clear and consistent evidence that HCW who had or were at risk for contact with infected patients, were more likely to experience psychological distress (Figure 3). Worry about becoming infected is a key stressor for HCW in the context of an outbreak as risk of infection has implications not only for their own health but also for that of their families (33). Evidence also indicated that being in quarantine contributes to distress, perhaps due to being isolated from the team (53), and that vicariously experiencing these risks can be detrimental for HCW mental health (25).

**Figure 2:**
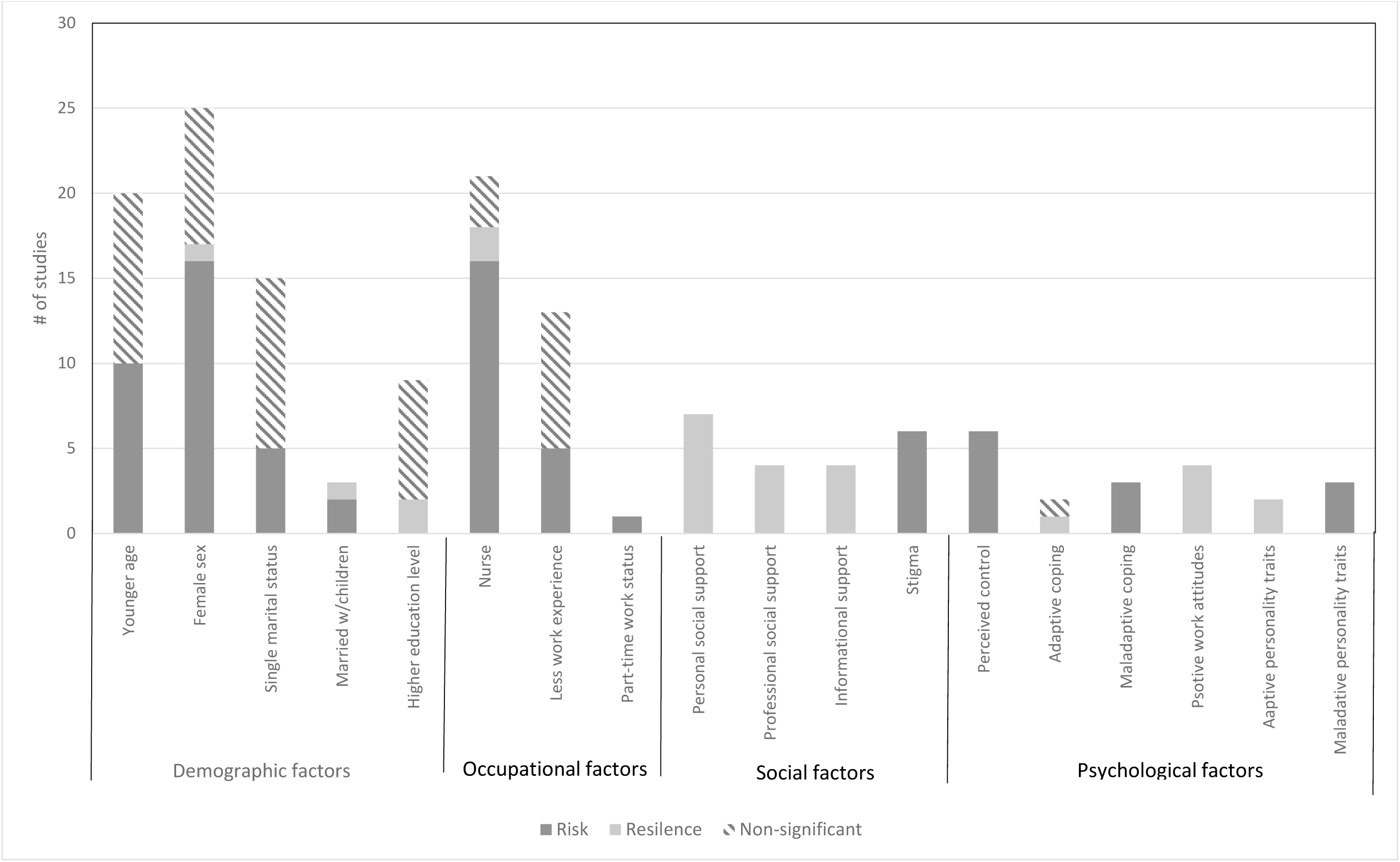
Findings from the studies that examined fixed (demographic and occupational) and modifiable (social and psychological) factors and associations with risk and resilience for psychological distress.

**Figure 3:**
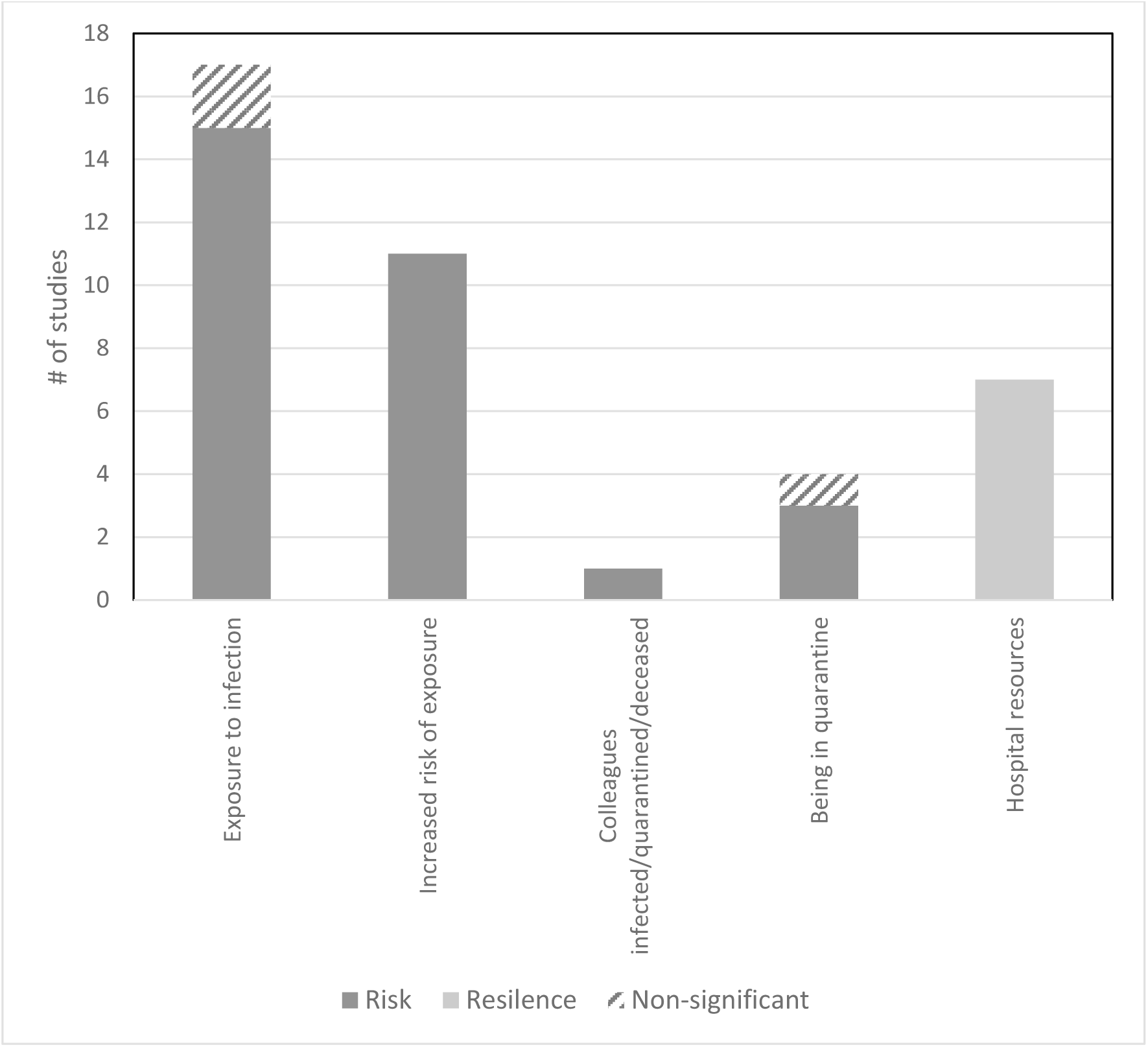
Findings from the studies that examined factors related to infection exposure and associations with risk and resilience for psychological distress.

Although modifiable factors were investigated by fewer studies (Figure 2), the evidence highlighted key areas to target to reduce HCW distress. Stigmatising attitudes from the public towards HCW were associated with greater distress. Public health campaigns that deliver accurate messages and highlight facts to reduce the fears underlying stigma (54), and counteract the fear-mongering cultivated through the media that can promote stigma during an infectious outbreak (55) could address this. Consistent with research on the stress-reducing effects of social support in general (56), and for HCW (57), the evidence indicated that perceiving social support was associated with lower distress. This can come from supervisors and co-workers (58), and through positive performance feedback (41). Such support can foster positive work attitudes and satisfaction (59), which were associated with lower distress. Evidence also indicated that harmful coping strategies, that may maintain or increase stress, were linked to greater distress. Interventions that target harmful coping are important for reducing distress, and other adverse health consequences, as HCW who experience post-traumatic stress during an outbreak and use harmful coping are at greater risk for substance abuse (60).

Perceptions of control were consistently associated with lower distress in the evidence reviewed. Although feeling a loss of control may be inevitable during an infectious outbreak, as perceptions of risk are inversely related to perceived control (61), efforts focused on increasing a sense of autonomy can be effective for reducing distress in HCW during stressful times (62). The evidence reviewed suggests that this might be accomplished by providing HCW with the resources needed to manage the risk of infection, such as through personal protective equipment, adequate training, and clear guidelines, information, and protocols for infection control, which were each linked to lower distress. This is consistent with research that found that access to information and provision of needed resources increased a sense of empowerment among ICU nurses (63).

### Limitations and strengths

There are several limitations of this rapid systematic review. Conducting the review during the ongoing outbreak of COVID-19 imposed time constraints. This meant that a formal quality appraisal was not conducted, though only studies with analytic samples over 80 were included as a proxy measure of quality control. Most study samples were quite large, increasing confidence in the generalisability of the findings. Also, we only included published peer-reviewed literature and did not search more thoroughly through grey literature or online pre-print repositories. In terms of the evidence base, the majority of the studies were cross-sectional, providing only a snapshot of the factors associated with HCW’s psychological distress. This limits conclusions about the direction of causality between the factors and distress, especially for those that are modifiable. Only three studies examined the potential long-term effects of the risk and resilience factors on HCW’s mental health by using follow-up and time-lagged designs (14, 21, 51), providing some support for the assumed effect of the factors on distress. More research is needed to track the associations of risk/resilience factors over time with distress and the extent to which certain factors are linked to sustained or transient distress. Although a number of studies investigated fixed factors and infection-related factors, there were relatively fewer studies that examined how modifiable factors were linked to distress (Figures 2-3). More research focusing on these factors is needed to provide a more solid evidence base about potential targets for clinical intervention and treatment. Several studies used unvalidated measures of psychological distress, raising concerns about whether the findings would be the same had validated measures been used. Few studies considered potential confounders in the associations with distress, compared found associations in matched non-HCW samples, or the extent to which the factors were predictive of distress outside of an outbreak. These limitations may have contributed to the equivocal findings noted for several of the factors reviewed.

These limitations are balanced by several strengths of the Review. Conceptually organising the factors according to risk or resilience and further as to whether they were fixed or modifiable provided a framework for identifying who might be at most risk for psychological distress to facilitate appropriate clinical intervention, and for noting which factors would be suitable targets for appropriate interventions. The Review included evidence from across several infectious disease outbreaks, increasing the possibility that the risk and resilience factors identified can be more generally applicable across different outbreaks. Lastly, conducting a series of search updates ensured integration of the most recent evidence from the ongoing COVID-19 outbreak into the review at the time of submission.

### Implications

Whereas other reviews have documented the extent of distress experienced by HCW during an outbreak (2), the current review highlights the profiles of HCW most at risk for psychological distress and psychiatric morbidity during an outbreak and identified modifiable factors that warrant further investigation as possible points of intervention to mitigate distress. Further research focusing on possible interactions among these factors would be useful to gain a better understanding of both the risk profiles and key modifiable factors, as the evidence reviewed did not consistently examine this.

Because there is evidence that the psychological distress from working during an outbreak can persist for two to three years after the outbreak (45, 51, 64), monitoring and providing appropriate support should continue beyond the outbreak period to ensure mental health recovery, especially among HCW who are most at risk. Our findings suggest that particular attention should be paid to female HCW and nurses (regardless of sex), and those who come in contact with infected patients or their environments to ensure that they receive necessary resources and provision of support to manage psychological distress. Proactive approaches at the organisational level can be effective (58) and may be necessary, as a study of HCW during the COVID-19 outbreak in China found that mental health resources and services were mainly used by those experiencing mild and subthreshold levels of psychological distress rather than those who experienced more severe distress (11). Evidence from randomised controlled trials suggests that third-wave cognitive behavioural therapeutic approaches, such as mindfulness (65), gratitude (66), and self-compassion (67), are effective for reducing stress and burnout among healthcare professionals, and could be beneficial. In low-resource settings, peer-support is one option that has been shown to be effective for reducing occupational distress in HCW (58). Raising awareness of the impact of an infectious outbreak on HCW mental health, providing appropriate treatment and therapy, and fostering proactive approaches such as an organisational culture of support, are suggested as possible approaches that can help prepare HCW for future outbreaks and address any persistent, long-term distress following the outbreak.

## Data Availability

Protocol for the systematic review is available on PROSPERO: https://www.crd.york.ac.uk/PROSPERO/; registration ID: CRD42020178185

## Author Contributions

- FS and JO designed and conceived the study. JO and FS searched for evidence. JO and FS screened and analysed the data
- FS drafted the work. JO and FS revised it critically for intellectual content
- FS and JO approved the final piece of work for publication
- FS and JO agree to be accountable for the work

